# Results from the first South African Health Products Regulatory Authority-approved randomised trial evaluating supplementation of standard antibiotic therapy with a commercially available probiotic in South African women with bacterial vaginosis

**DOI:** 10.1101/2020.05.04.20090282

**Authors:** Anna-Ursula Happel, Ravesh Singh, Nireshni Mitchev, Koleka Mlisana, Heather B. Jaspan, Shaun L. Barnabas, Jo-Ann S. Passmore

## Abstract

Abstract

**Background:** Bacterial vaginosis (BV) increases HIV risk and adverse reproductive outcomes in women. The standard-of-care (SOC) for BV is antibiotic treatment; however, cure rates are low and recurrence frequent. In South Africa, no trial comparing probiotics to SOC for BV has been approved by the South African Health Products regulatory Authority (SAHPRA). We aimed to explore the South African regulatory and ethics environment to evaluate adjunctive probiotics for improvement of BV treatment in a randomized single-blinded trial of a locally sourced oral-vaginal-combination probiotic for vaginal health.

**Methods:** South African women with symptomatic vaginal discharge were screened for BV and sexually transmitted infections (STIs) including *Trichomonas vaginalis, Mycoplasma genitalium, Neisseria gonorrhoae* and *Chlamydia trachomatis*. BV positive (by Nugent Scoring) STI negative women were randomized to Metrogel™ alone (n=12) or Metrogel™ followed by a commercially available South African oral/vaginal probiotic (Vagiforte PLUS^®^ Combo Pack; n=18). BV cure at one month was the primary endpoint. Secondary endpoints were recurrence, symptoms, vaginal microbiota and genital inflammation over five months post-treatment, and acceptability of the administered probiotic.

**Results:** SAHPRA reviewed and acknowledged this trial. Overall, 44.8% of women cleared BV one month post-treatment. Despite confirmed viability of probiotic species contained in the commercial oral/vaginal probiotic, they did not appear to colonize the female genital tract of most women in the intervention group. No significant differences in BV cure rates, vaginal pH, microbiota nor IL-1α concentrations were found between SOC and intervention groups, although we were underpowered to detect small differences. Acceptability and adherence to the probiotic product was high.

**Conclusion:** Navigation of the SAHPRA registration process for evaluating a commercial probiotic in a randomised trial has laid the path for future trials of improved probiotic products for vaginal health in South Africa with adequate sample sizes. Acceptability of vaginally applied probiotics was high in South African women. Improvements in both the content and delivery of commercially available probiotic products for vaginal health should be considered.

**Trial registration:** This trial was registered on 17 October 2017 with the South African National Clinical Trial Register of the Department of Health (DOH-27-1117-5579).

## Background

Globally, bacterial vaginosis (BV), a polymicrobial dysbiosis of the vagina, is the most common genital condition of women of reproductive age (1), and increases risk of adverse pregnancy outcomes (2,3) and acquisition and transmission of sexually-transmitted infections (STIs), including HIV (4,5), possibly due to the associated genital inflammation (6). Antibiotics such as metronidazole and clindamycin remain the standard of care (SOC) for treating BV, although it is estimated that more than 50% of women experience recurrent episodes within 6-12 months (7,8). Most countries treat BV as part of syndromic management (9), although >85% of BV cases are asymptomatic while still being associated with significantly elevated genital inflammatory cytokine profiles (10). In Africa, where epidemics of BV, STIs and HIV converge (11) and genital inflammation associated with even asymptomatic BV may increase HIV risk (5,12), an urgent need to rethink and improve the SOC for treating BV exists.

Several clinical studies have evaluated *Lactobacillus-containing* probiotics as an adjunct to antibiotics in treating BV (13,14). However, there is clinical equipoise as to whether adjunctive probiotics improve BV cure and/or recurrence rates, as a recent meta-analysis (15) concluded that adjunctive probiotics may improve BV cure rates but that there is currently not enough high-quality clinical evidence to support this approach as superior to antibiotic treatment alone. While randomized controlled trials (RCTs) assessing the effects of probiotics on BV management have been performed in several African countries (16,17), only one exploratory pilot study has recently been performed in South Africa (18). As the microbial composition of the lower female genital tract (FGT) in health and dysbiosis may be influenced by regional factors, including diet, vaginal insertion, hygiene practices and possibly host genetics (19–21), it is important to conduct such trials in South African women to evaluate the effectiveness of probiotics for vaginal health in a local context.

Recently, we surveyed >100 commercially-available probiotics currently on the market in South Africa as over-the-counter (OTC) products, of which only four were indicated for vaginal health and only one was intended for vaginal application (22). Although in South Africa the registration of medicines, probiotics and health supplements along with their use in clinical trials is regulated by the South African Health Product Regulatory Authority (SAHPRA), none of these OTC probiotics had been formally reviewed by SAHPRA and no trial evaluating the impact of probiotics on health outcomes had previously been SAHPRA-approved or -acknowledged.

We therefore aimed to test the South African regulatory environment for evaluating vaginal probiotics in clinical trials using a locally sourced OTC product – Vagiforte^®^ PLUS Combo Pack. This probiotic was the only local product administered orally as well as vaginally and was intended for a prolonged treatment duration of 15 days. This single-blinded, randomized trial enrolled symptomatic BV positive South African women to compare vaginal metronidazole gel treatment alone (SOC) to a combination of metronidazole and the locally manufactured oral/vaginal probiotic, with BV cure being the primary endpoint. Secondary endpoints included adherence, acceptability and preference of the probiotic, and the changes in vaginal pH, symptoms and vaginal cytokine concentrations (interleukin [IL]-1α), as well as durability of any effect three and five months post-treatment. Changes in the vaginal concentrations of beneficial *Lactobacillus* spp. and non-optimal bacterial spp. (including *Gardnerella vaginalis, Prevotella bivia*, Bacterial Vaginosis-Associated Bacterium 2 (BVAB2), *Megasphaera* 1 and *Atopobium vaginae)* and evidence of vaginal colonization of the bacterial species contained in the administered probiotic were also evaluated. This first SAHPRA-registered probiotic trial intended to lay the foundation for future testing of novel or optimized probiotic products for BV management in South African women in RCTs with adequate sample sizes.

## Methods

### Regulatory approval

Prior to conducting this trial, discussions with SAHPRA authorities were initiated in March 2016 and key areas that needed addressing were identified, including a decision on whether a notification or full application for the trial was necessary. Typically, the regulator requires full applications for products not registered in South Africa; or for those not being used for their registered indication, dose, or formulation. Alternatively, SAHPRA notification is required for phase IV clinical studies of an approved medication within its approved dosage, formulation and indication. SAHPRA confirmed that the trial did not require a full application as the product was already available OTC in South Africa, thus a notification application was submitted to SAHPRA and approved (SAHRPA Ref 20161201; PI: S. Barnabas).

### Eligibility criteria

Women were recruited from a South African public sector STI clinic (Spencer Road Clinic) and from the UCT Student Wellness Centre in Cape Town, South Africa. Eligible women were 18-45 years old and seeking care for vaginal discharge. All eligible women were tested for BV (by Nugent scoring) and STIs, including *C. trachomatis, N. gonorrhoea, T. vaginalis*, and *M. genitalium* by TaqMan® Assays (Fast Track Diagnostics). Inclusion criteria were being BV positive (Nugent 7-10) but negative for any STI. Exclusion criteria included being pregnant, breastfeeding, pelvic inflammatory disease, living with HIV, having a known allergy to metronidazole, and/or currently using any other antibiotics or natural remedies in the urogenital area. Women who acquired an STI or had recurrent BV over the course of the trial were referred for treatment but not excluded, and any concomitant medication (including antibiotics) taken over the course of the study was recorded. Study visits were planned such that women were not menstruating nor reported having unprotected sex or douching in the 48 hours prior to sampling. All women were tested for HIV (Rapid Anti-HIV (1&2) test; InTec products, Inc., China) and pregnancy (hCG Pregnancy test; Homemed™, South Africa) at screening.

### Randomisation and blinding

Randomisation was performed using the pseudorandom number generator in Microsoft Excel 2016 (MT19937) by research pharmacists at the UCT CRC who were not involved with clinical procedures and/or screening processes and only dispensed the investigational product. Researchers and laboratory staff involved in sample and data analysis were blinded to the randomisation process. The research nurse who conducted the clinic visits was not blinded as she interacted directly with participants. After the database lock and a primary blinded analysis, the unblinded treatment allocations were released.

### Dosing regimens

Eligible women either received topical metronidazole only (0.75% gel, 5 g vaginally, once a day for 5 days; MetroGel™ V, iNova Pharmaceuticals, South Africa, SOC group) or topical metronidazole followed by a 15-day treatment course of Vagiforte® PLUS Combo Pack (Bioflora, South Africa, Lot # 21918; intervention group), which included five days of oral probiotic capsules followed by ten days of oral capsules together with twice daily vaginal spray. Vagiforte® PLUS oral capsules and each metered dose of the vaginal spray contained lyophilized *L. acidophilus, L. rhamnosus GG, B. bifidum* and *B. longum* at ≥ 2×10^9^ colony-forming units (CFU).

### Laboratory quality control of Vagiforte® PLUS lot used in the study

The contents and concentrations of each microbial species of Vagiforte^®^ PLUS Combo Pack Lot #21918 were confirmed prior to initiation of the trial (Additional Figure 1). Briefly, one full oral or vaginal dose was dissolved in Brain Heart Infusion broth (supplemented with 0.1% starch and 1% yeast, sBHI), serially diluted and plated in triplicates onto sBHI agar plates. Plates were incubated at 37°C for 48 hours under anaerobic conditions (using Oxoid™ AnaeroGen™ 2.5L Sachets, Thermo Fisher Scientific Inc., USA). The CFU per well were counted and the average concentration per dose was calculated. Contents were confirmed by MALDI-TOF (MALDI Biotyper, Bruker Daltonik, USA).

**Additional Figure 1.**
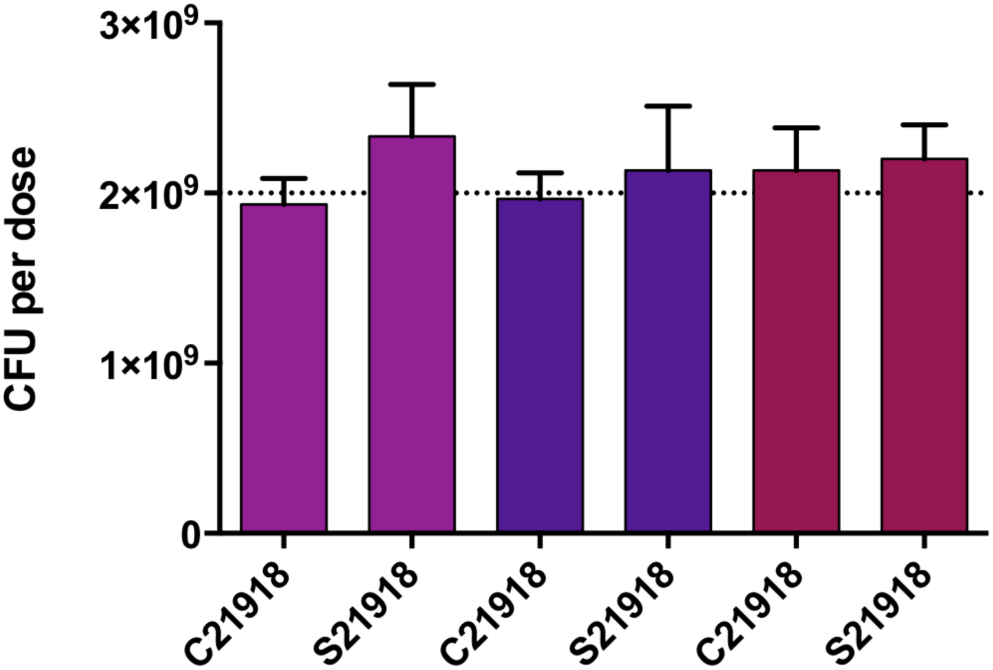
Bacterial concentration per dose unit of Vagiforte® PLUS Combo Pack. The CFU per dose of three boxes (pink, purple and red) of Vagiforte® Plus Combo Pack lot #21918 containing oral capsules (C) and vaginal spray (S) was determined using serial dilutions and compared to the manufacturers claim of 2×109 CFU per dose (dotted line).

### Clinical procedures and sample collection

At screening, a vulvo-vaginal swab for STI testing and a posterior fornix and lateral wall swab to screen for BV by Nugent Scoring were collected. At enrolment (pre-treatment), and one, three and five months post-treatment, genital samples were collected in the following order under speculum examination: (1) a vulvo-vaginal swab for STI testing; (2) a posterior fornix and lateral wall swab to screen for BV (by Nugent Scoring) and to measure vaginal pH (using a colour-fixed indictor pH strip; Macherey Nagel); and (3–4) two lateral wall swabs to measure genital IL-1α (as marker of genital inflammation, by ELISA) and vaginal microbiota (by qPCR). In addition, women completed a questionnaire on demographics, reproductive health and sexual behaviour at enrolment, and a questionnaire assessing feasibility, acceptability and adherence to the administered products, vaginal symptoms and adverse events at follow up visits. Product preference was assessed at the final visit by questionnaire. Adherence was measured by self-report in medication diaries and questionnaires, as well as return of empty packages at the one-month follow-up visit.

### Testing for STIs and BV

The commercial TaqMan® FTD STD9 (Fast Track Diagnostics, Luxembourg) kit, performed as per manufacturer’s instructions, was used to test for *N. gonorrhoeae, C. trachomatis, T. vaginalis* and *M. genitalium*. As positive controls, genomic DNA extracts from the following ATCC^®^ strains were included: *N. gonorrhoeae* (ATCC® 700825), *C. trachomatis* (ATCC® VR-885), *T. vaginalis* (ATCC® 30001) and *M. genitalium* (ATCC® 33530). BV was diagnosed by Gram staining of vaginal smears and Nugent Scoring. Slides were assessed microscopically and assigned a score between 0 and 10, with a score of 0-3 considered BV negative, 4-6 intermediate microbiota, and 7-10 BV positive.

### Quantitative measurement of vaginal bacterial concentrations

To assess changes in the vaginal microbiota, vaginal concentrations of common vaginal *Lactobacillus* spp. (including *L. crispatus, L. jensenii, L. iners, L. gasseri, L. vaginalis, L. mucosae)*, BV-associated organisms (including *G. vaginalis, P. bivia*, BVAB2, *Megasphaera* 1 and *A. vaginae*), and bacterial species contained in the administered probiotic product (including *L. acidophilus, L. rhamnosus, B. bifidum* and *B. longum*) were assessed using commercially available Applied Biosystems™ TaqMan® Assays (Thermo Fisher Scientific Inc., USA; Assay IDs Ba04646245_s1, Ba04646258_s1, Ba04646257_s1, Ba04646234_s1 for *Lactobacillus* spp. and Ba04646236_s1, Ba04646278_s1, Ba04646229_s1, Ba04646230_s1, Ba04646222_s1 for BV-associated species). For *L. vaginalis* (KF875988.1) and *L. mucoasae* (NR_024994.1) Thermo Fisher Scientific designed custom probes based on the referenced nucleotide sequence. The TaqMan probe sequences for *L. acidophilus* and *L. rhamnosus* (Haarman and Knol, 2006), as well as *B. bifidum* and *B. longum* (Haarman and Knol, 2005) were previously published. Using an ABI 7500 Real-Time PCR Detection System (Thermo Fisher Scientific Inc., USA) we quantified each of the targets. The quantification was performed using amplicons generated from plasmids for each of the targets (TaqMan™ Vaginal Microbiota Extraction Control). Serial dilutions, ranging from 10^0^ to 10^9^ molecules per µl of sample were used to generate standard curves. Bacterial concentrations were normalized to 16S rRNA gene concentrations, as recommended by the manufacturer.

### Measurement of IL-1α concentrations

Samples were thawed on ice and filtered by centrifuging at 1950 g for 10 minutes at 4°C in SPIN-X® 0.2µM cellulose acetate filters to exclude mucus and debris prior to performing the ELISA. Human IL-1α concentrations were measured using a commercial ELISA (E-EL-H008, Elabscience®, USA), according to the manufacturer’s instructions. IL-1α concentrations were calculated based on the standard curve, using standards provided with the kit. The detection range was 1.25-125 pg/mL, and all values below the detection limit were recorded as half of the lowest concentration measured.

### Statistical analyses

As this was a pilot trial intended to test the regulatory environment in South Africa for conducting a trial assessing probiotics, it was not powered for efficacy (defined as BV cure). None-the-less, GraphPad Prism6® (GraphPad Software, USA), STATA version 11.0 (StataCorp, USA) and R were used for statistical analyses and to generate graphs. Mann-Whitney U tests were used to compare groups of continuous variables and two-sided Fisher’s exact tests were used for categorical variables. 95% confidence intervals and p-values ≤0.05 were considered significant.

## Results

### SAHPRA approval

This trial intended to explore the regulatory environment in South Africa governing probiotic trials, in order to lay the foundation for future trials of novel probiotic products under development. SAHPRA did not require a full application but rather a notification, as the probiotic product was not currently considered/regulated as a medicine but as a health supplement, and available OTC in South Africa.

### Cohort behavioural and biomedical characteristics

A total of 96 women seeking care for vaginal discharge were screened for eligibility (Figure 1), of which the majority (n=90) were recruited via the UCT Student Wellness Centre. One woman tested HIV positive at screening, and thus was excluded and referred for care. Of those screened, 43 women (45.3%) were confirmed to have BV by Nugent Scoring (Nugent 7-10; Table 1), confirming that symptomatic vaginal discharge is a very imprecise tool for predicting the presence of BV (11), with a positive predictive value of only 45.3% in this cohort. In addition, 17.9 % (17/95) had an STI, with *T. vaginalis* in 9.5% (9/95) and *C. trachomatis* in 8.4% (8/95) being the most common STIs detected. Of the 43 BV positive women, 33/43 were eligible for randomization (BV+ but STI-).

**Figure 1.**
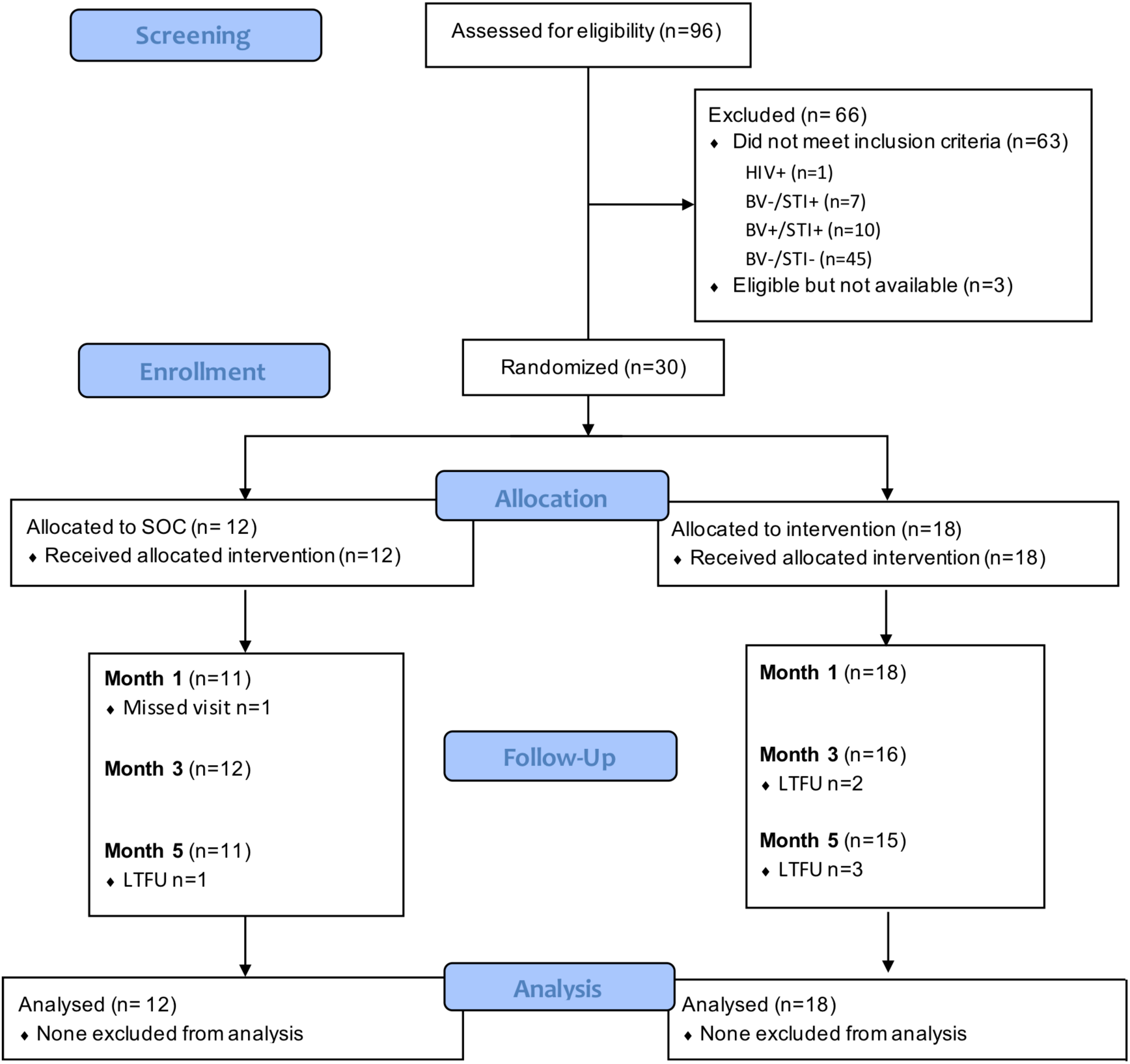
Consort diagram. A total of 96 women with symptomatic discharge were screened for eligibility Participants were tested for BV by Nugent Scoring on for STIs (including *C. trachomatis, T. vaginalis, M. genitalium* and *A. vaginae)* by Multiplex PCR. Eligible participants (BV positive but STI negative) were randomly assigned to the SOC (Metrogel™ only) or intervention arm (Metrogel™ plus Vagiforte® Plus Combo Pack). Follow-up visits took place 1, 3 and 5 months post-treatment.

**Table 1.** Laboratory-diagnosed BV and STIs at screening.

| **Characteristic** |  |
| --- | --- |
| **Age participant [years, median (IQR)]** | 22 (20-25) |
| **Self-identified race [% (n/N)]** |  |
| Black | 67.7 (65/96) |
| Coloured | 19.8 (19/96) |
| White | 12.5 (12/96) |
| **Bacterial vaginosis [% (n/N)]<sup>a</sup>** |  |
| Positive (Nugent Score 7-10) | 45.3 (43/95) |
| Intermediate (Nugent Score 4-6) | 15.8 (15/95) |
| **Bacterial and protozoal infections [% (n/N)]<sup>a</sup>** |  |
| *T. vaginalis* | 9.5 (9/95) |
| *C. trachomatis* | 8.4 (8/95) |
| *M. genitalium* | 3.2 (3/95) |
| *N. gonorrhoea* | 2.1 (2/95) |
| **Multiple conditions [% (n/N)]<sup>a</sup>** |  |
| BV+/STI- | 34.7 (33/95) |
| BV-/STI+ | 7.4 (7/95) |
| BV+/STI+ | 10.5 (10/95) |
| BV-/STI- | 47.4 (45/95) |
| Multiple STIs | 4.2 (4/95) |

<sup>a</sup>One participant tested HIV positive, thus screening was aborted.

Of the 33 women who were eligible, three were no longer interested in participating, 12 were randomized to the SOC arm (vaginal MetroGel™ alone; 5 days), while 18 were randomized to the intervention arm (vaginal MetroGel™ 5 days, followed by 15 days of Vagiforte®). Randomised women were a median age of 22 years (IQR 20-26 years) old, predominantly single (26/30, 86.7%), and self-identified as black (20/30, 66.7%). About one-third reported being previously pregnant (8/30), and one-third reported currently using hormonal contraception (11/30; Table 2). Their median age at sexual debut was reported to be 17 years (IQR 16-19) with a median of five lifetime sexual partners (IQR 3-8). Reporting of oral sex was common (21/30, 70.0%), while reporting of anal sex was less common (2/30, 6.7%). About half reported regular condom use, and very few (n=4) reported previously being diagnosed or treated for an STI (Table 2. Almost half reported that they were currently smoking, a factor that has previously been associated with the risk of BV (25–27). Almost all enrolled women (28/30; 93.3%) reported a history of chronic vaginal discharge. About a third reported using their fingers and water to clean their vaginas internally, while other vaginal practices (such as cleansing with cloth or using traditional South African herbs) were reported less commonly. A third of women reported believing that using vaginal products (either commercial and traditional) to treat their vaginal discharge was beneficial for their vaginal health, and an equal proportion reported concerns that these vaginal products were causing their vaginal discharge and malodour (Table 2). More than half reported previous use of prescription medicine to reduce vaginal discharge and malodour, supporting the history of long-standing vaginal discharge and/or infections in these women. More than half used tampons while menstruating, suggesting that they may be likely to be comfortable with the vaginal application of the probiotic (Table 2). None of these characteristics evaluated differed by study arm.

**Table 2.**
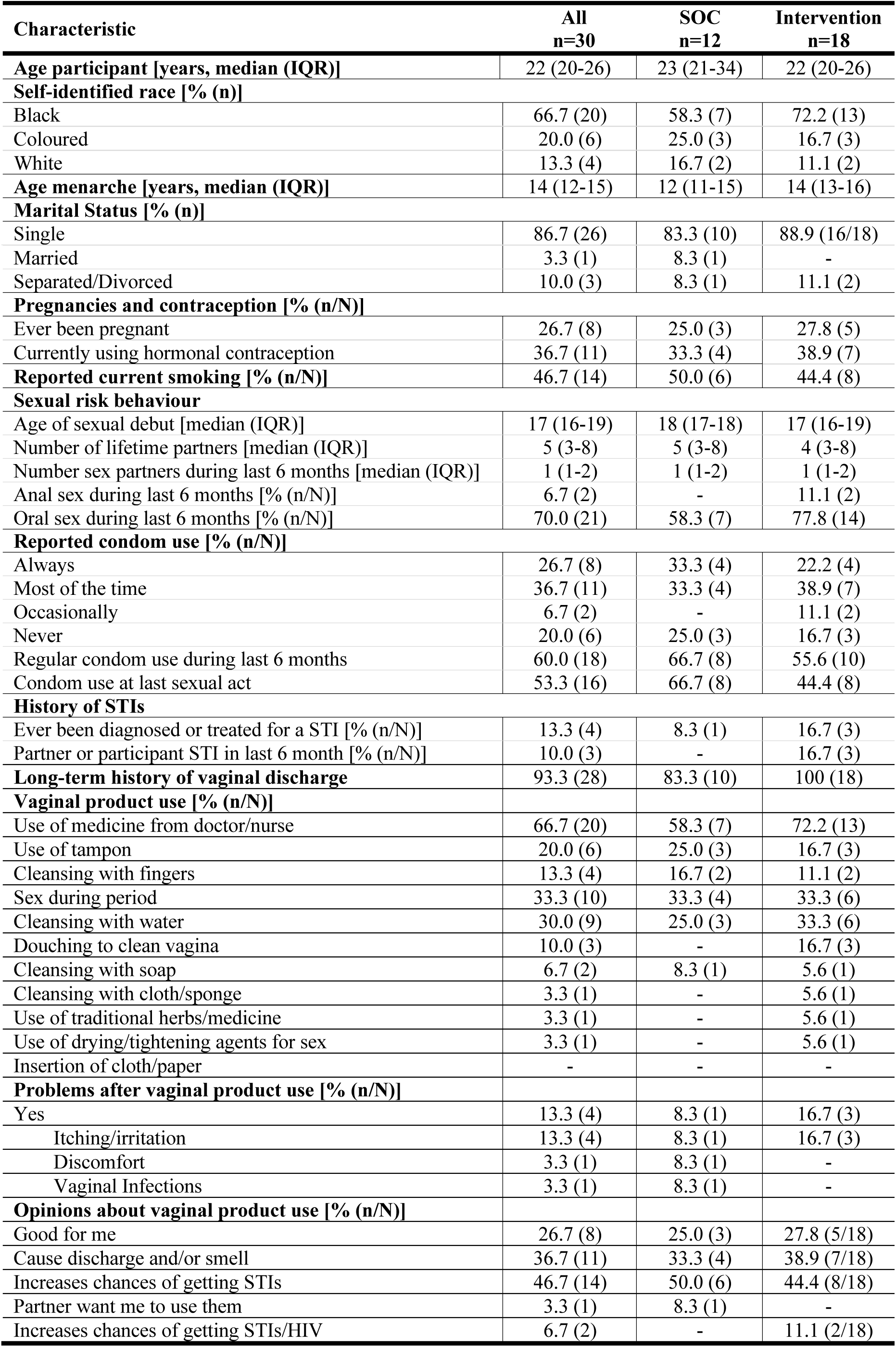
Cohort characteristics at enrolment.

| Characteristic | All<br>n=30 | SOC<br>n=12 | Intervention<br>n=18 |
| --- | --- | --- | --- |
| **Age participant [years, median (IQR)]** | 22 (20-26) | 23 (21-34) | 22 (20-26) |
| **Self-identified race [% (n)]** |  |  |  |
| Black | 66.7 (20) | 58.3 (7) | 72.2 (13) |
| Coloured | 20.0 (6) | 25.0 (3) | 16.7 (3) |
| White | 13.3 (4) | 16.7 (2) | 11.1 (2) |
| **Age menarche [years, median (IQR)]** | 14 (12-15) | 12 (11-15) | 14 (13-16) |
| **Marital Status [% (n)]** |  |  |  |
| Single | 86.7 (26) | 83.3 (10) | 88.9 (16/18) |
| Married | 3.3 (1) | 8.3 (1) | - |
| Separated/Divorced | 10.0 (3) | 8.3 (1) | 11.1 (2) |
| **Pregnancies and contraception [% (n/N)]** |  |  |  |
| Ever been pregnant | 26.7 (8) | 25.0 (3) | 27.8 (5) |
| Currently using hormonal contraception | 36.7 (11) | 33.3 (4) | 38.9 (7) |
| **Reported current smoking [% (n/N)]** | 46.7 (14) | 50.0 (6) | 44.4 (8) |
| **Sexual risk behaviour** |  |  |  |
| Age of sexual debut [median (IQR)] | 17 (16-19) | 18 (17-18) | 17 (16-19) |
| Number of lifetime partners [median (IQR)] | 5 (3-8) | 5 (3-8) | 4 (3-8) |
| Number sex partners during last 6 months [median (IQR)] | 1 (1-2) | 1 (1-2) | 1 (1-2) |
| Anal sex during last 6 months [% (n/N)] | 6.7 (2) | - | 11.1 (2) |
| Oral sex during last 6 months [% (n/N)] | 70.0 (21) | 58.3 (7) | 77.8 (14) |
| **Reported condom use [% (n/N)]** |  |  |  |
| Always | 26.7 (8) | 33.3 (4) | 22.2 (4) |
| Most of the time | 36.7 (11) | 33.3 (4) | 38.9 (7) |
| Occasionally | 6.7 (2) | - | 11.1 (2) |
| Never | 20.0 (6) | 25.0 (3) | 16.7 (3) |
| Regular condom use during last 6 months | 60.0 (18) | 66.7 (8) | 55.6 (10) |
| Condom use at last sexual act | 53.3 (16) | 66.7 (8) | 44.4 (8) |
| **History of STIs** |  |  |  |
| Ever been diagnosed or treated for a STI [% (n/N)] | 13.3 (4) | 8.3 (1) | 16.7 (3) |
| Partner or participant STI in last 6 month [% (n/N)] | 10.0 (3) | - | 16.7 (3) |
| **Long-term history of vaginal discharge** | 93.3 (28) | 83.3 (10) | 100 (18) |
| **Vaginal product use [% (n/N)]** |  |  |  |
| Use of medicine from doctor/nurse | 66.7 (20) | 58.3 (7) | 72.2 (13) |
| Use of tampon | 20.0 (6) | 25.0 (3) | 16.7 (3) |
| Cleansing with fingers | 13.3 (4) | 16.7 (2) | 11.1 (2) |
| Sex during period | 33.3 (10) | 33.3 (4) | 33.3 (6) |
| Cleansing with water | 30.0 (9) | 25.0 (3) | 33.3 (6) |
| Douching to clean vagina | 10.0 (3) | - | 16.7 (3) |
| Cleansing with soap | 6.7 (2) | 8.3 (1) | 5.6 (1) |
| Cleansing with cloth/sponge | 3.3 (1) | - | 5.6 (1) |
| Use of traditional herbs/medicine | 3.3 (1) | - | 5.6 (1) |
| Use of drying/tightening agents for sex | 3.3 (1) | - | 5.6 (1) |
| Insertion of cloth/paper | - | - | - |
| **Problems after vaginal product use [% (n/N)]** |  |  |  |
| Yes | 13.3 (4) | 8.3 (1) | 16.7 (3) |
| Itching/irritation | 13.3 (4) | 8.3 (1) | 16.7 (3) |
| Discomfort | 3.3 (1) | 8.3 (1) | - |
| Vaginal Infections | 3.3 (1) | 8.3 (1) | - |
| **Opinions about vaginal product use [% (n/N)]** |  |  |  |
| Good for me | 26.7 (8) | 25.0 (3) | 27.8 (5/18) |
| Cause discharge and/or smell | 36.7 (11) | 33.3 (4) | 38.9 (7/18) |
| Increases chances of getting STIs | 46.7 (14) | 50.0 (6) | 44.4 (8/18) |
| Partner want me to use them | 3.3 (1) | 8.3 (1) | - |
| Increases chances of getting STIs/HIV | 6.7 (2) | - | 11.1 (2/18) |

### Adherence to and safety of MetroGel™ and the oral/vaginal probiotic

Women attended a total of 143 visits, with 25/30 women completing all visits (Figure 1). All women (n=30) reported completing the entire five-day course of MetroGel™. Similarly, all women randomised to the intervention group (n=18) reported completing the course of oral probiotic capsules, and the majority (n=16) also reported completing the vaginal probiotic spray. Overall, a total of 110 adverse events (AEs) occurred over the course of the trial, of which most (76.4 %; 84/110) were considered not to be related to study products. Most AEs were mild (grade 1-2, 109/110, 99.1%) and only one non-related AE was grade 3. Unrelated AEs included: headaches (20 periods of headache reported by 5 women), flu-like symptoms (n=18, reported by 12 women), acquisition of any STI [including *C. trachomatis* (n=8), *T. vaginalis* (n=7), *M. genitalium* (n=5) and *N. gonorrhoea* (n=1)], menstrual pain (n=7), gastro-intestinal complaints (n=5), temporary vaginal discomfort related to vaginal intercourse (n=7), anaemia (n=2) and cat allergy (n=1), buttock pain (n=1), and anxiety (n=1). More than half of the women (53.3%; 16/30) acquired an STI over the course of the study.

Of those AEs that were considered to be related to the use of the study products (n=26), the majority was associated with vaginal MetroGel™ use and reported by women who were randomised to the SOC arm (80.8%; 21/26). Overall, more than half of the participants (17/30; 56.7 %) reported experiencing genital AEs, including vaginal itching/irritation (9/30; 30.0%), candida infections (5/30; 16.7%), discomfort (3/30; 10.0%), increased discharge (2/30; 6.7%), spotting (1/30; 3.3%), or constipation after MetroGel™ use (1/30; 3.3%). Vaginal candida infection and constipation are published side effects of MetroGel™, and vaginal itching, discomfort and discharge are likely to be symptoms of candidiasis (28). One third of women (5/18; 27.8%) reported AEs which may have been associated with probiotic use, including vaginal itching/discomfort (2/18; 11.1%), increased vaginal discharge (1/18; 5.6%), increased nipple sensitivity (1/18; 5.6%), and nausea (1/18; 5.6%). Overall, the adherence to both products was high and generally associated with a good safety profile.

### Acceptability of the oral-vaginal combination probiotic in South African women

At the final visit, women in the intervention arm were asked by the research nurse to complete a questionnaire to evaluate the acceptability of the probiotic product used during the trial. The majority of women (9/15, 60.0%) reported that they liked using the combination probiotic, because it was easy to use or resulted in improved vaginal symptoms (Figure 2A). Most women (12/15, 80.0%) reported that they preferred the oral capsule over the vaginal spray application (Figure 2B), as the spray was difficult to use, “messy and smelly” or AEs occurred. Despite seven women reporting that they did not like using the oral/vaginal probiotic, the vast majority (14/15, 93.3%) believed that they received some benefit from this product and would buy it to use it again, primarily to prevent rather than to treat BV (Figure 2C). The majority (13/15, 86.7%) also said they would recommend it to other women. In terms of future product design, women reported that they would prefer a probiotic with oral application only rather than a combination of oral and vaginal or vaginal only administration (Figure 2D). If vaginal administration was required, women responded that they would prefer a tablet or gel rather than a tampon, spray or capsule. Women reported that they would much prefer to buy the probiotic at pharmacies (12/15, 80.0%) than health stores, a grocery store or clinic (Figure 2E). When questioned about where they prefer to seek advice regarding using probiotics for vaginal health, women responded that they would ask reproductive health nurses (13/15, 86.6%) and doctors (11/15; 73.3%) rather than pharmacists or get information from the internet (Figure 2F). When asked about how much they were willing to spend on such a product, none of the women responded that they would be willing to spend more than 200 South African rand (ZAR), which was less than the cost of the probiotic tested in this trial (costing ZAR 280 per treatment course).

**Figure 2.**
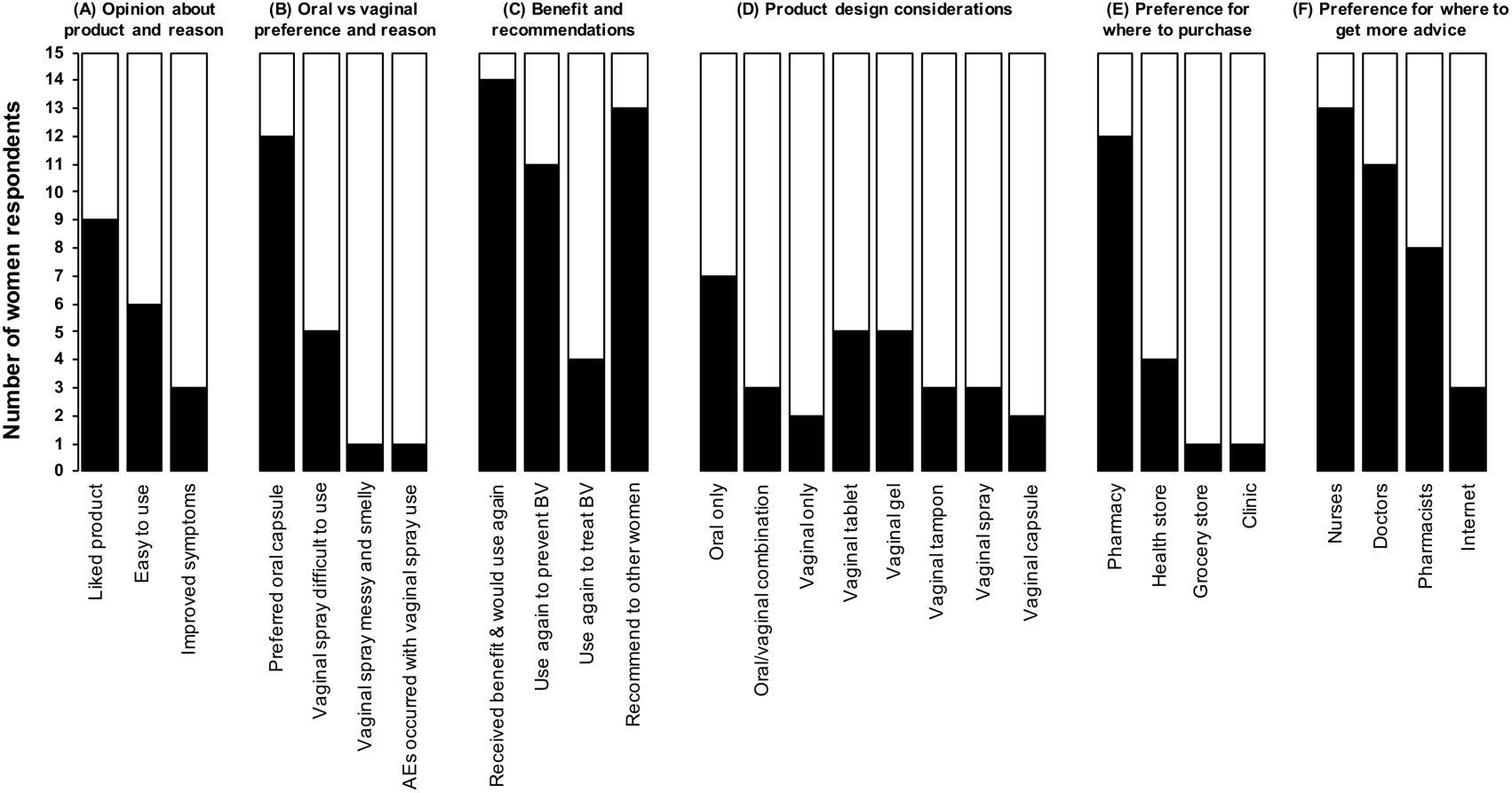
Acceptability and preference of probiotics for vaginal health. At the final visit, participants completed a questionnaire to assess their (A) opinion about the oral/vaginal probiotic they used during the trial, (B) why their preferred the oral or vaginal application, (C) their perceived benefits and whether they would use it again or recommend it to another woman, (D) their application preference for the development of a future probiotic for vaginal health, (E) where they would want to buy it and (F) from whom they would like to get advice regarding the use of probiotics for vaginal health.

### Comparing BV cure rates in SOC and intervention arms

Although our trial was not powered for efficacy, we were interested in an exploratory intention-to-treat (ITT) analysis to estimate possible benefits of administration of this locally sourced probiotic on BV cure and recurrence (Figure 3A). The overall BV cure rate (defined as achieving a Nugent score 0-3) was 44.8% (13/29) at month 1 (primary outcome), 46.4% (13/28) at month 3 and 53.8% (14/26) at month 5. At month 1, BV cure rates tended to be higher in the SOC arm (63.6 %; 7/11) than the intervention arm (33.3%; 6/18; p=0.113). However, almost half of the BV negative women in the SOC arm (3/7) subsequently re-tested BV positive (Nugent 7-10) at month 3, while more than half of the women in the intervention arm who were cured at month 1 (4/6) remained BV negative until the end of the trial. Vagiforte® PLUS did not appear to offer short-term benefits for BV cure compared to MetroGel™ alone, although it needs to be emphasised that this trial was not adequately powered.

**Figure 3.**
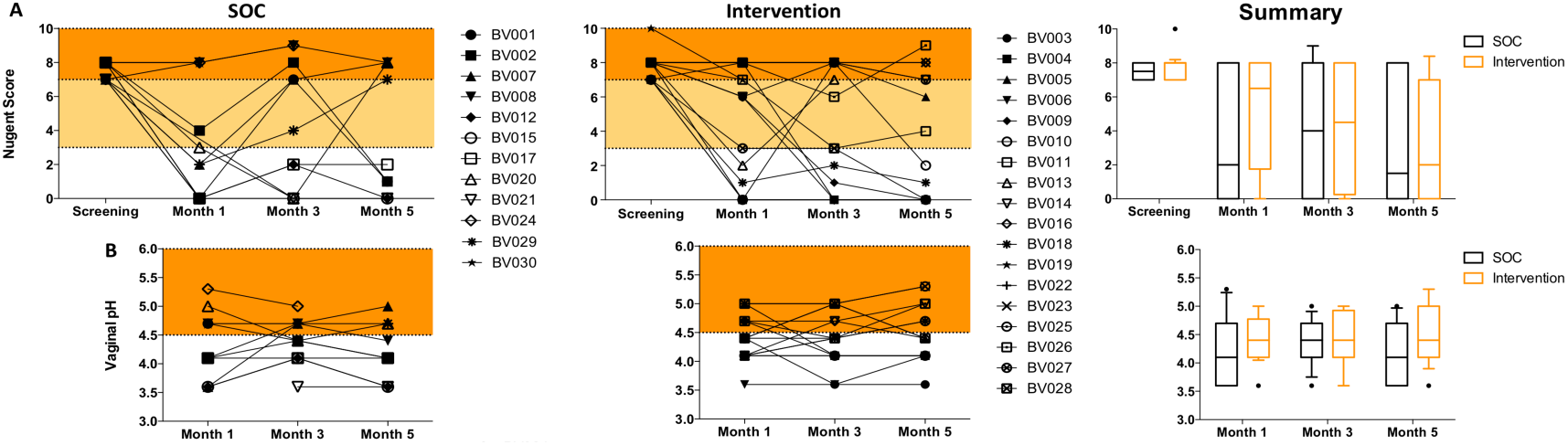
BV status and vaginal pH in the SOC and intervention group. (A) BV status was determined by Nugent scoring at screening, and one, three and five-months post-treatment. Nugent Score 0-3 = BV negative (white area); 4-6= BV intermediate (yellow) and 7-10=BV positive (orange). (B) Vaginal pH was measured using color-fixed indicator strips one, three and five-months post-treatment. A vaginal pH <4.5 (white area) is seen as protective. Each participant is represented by a symbol-coded dot, and the summaries show median and interquartile range.

### Comparing vaginal pH in SOC versus intervention arms

A pH>4.5 is one of the clinical criteria considered in diagnosing BV using Amsel’s Criteria (29) and a low vaginal pH is considered to be beneficial for vaginal health (Aroutcheva *et al*., 2001; Boskey *et al*., 1999). Thus, having a vaginal pH<4.5 after BV treatment was considered as a secondary endpoint for this study. In the SOC arm, 6/11 (54.5%) women had a vaginal pH<4.5 one month after treatment, compared to 11/18 (61.1%) in the intervention group (p=0.514; Figure 3B). Similarly, no significant difference between the SOC and intervention arms were noted at later time points. Nugent scores correlated significantly with vaginal pH (Spearman rho=0.71; p=0.0001).

### Comparing clinical symptoms in SOC versus intervention arms

The majority of women (24/29) reported an improvement of their vaginal symptoms one month after treatment, with similar rates in the SOC and intervention arms (Table 3). Of these, the majority (20/24) reported a decrease of vaginal discharge, and more than half reported a change in smell and/or colour of the discharge, independently of treatment arm. Similar improvements in vaginal symptoms were reported three months after treatment completion (Table 3).

**Table 3.** Reported vaginal symptoms one and three months after treatment completion.

| **Month 1 [% (n/N)]** | **Overall**<br>**n=29** | **SOC**<br>**n=11** | **Intervention**<br>**n=18** | **p-value<sup>a</sup>** |
| --- | --- | --- | --- | --- |
| Improvement of discharge | 82.8 (24/29) | 83.3 (10/11) | 77.8 (14/18) | 0.356 |
| Less discharge | 83.3 (20/24) | 90.0 (9/10) | 78.6 (11/14) | 0.437 |
| Change in colour | 54.2 (13/24) | 50.0 (5/10) | 57.1 (8/14) | 0.527 |
| Change in smell | 70.8 (17/24) | 70.0 (7/10) | 71.4 (10/14) | 0.643 |
| **Month 3 [% (n/N)]** | **n=28** | **n=12** | **n=16** |  |
| Improvement of discharge | 82.1 (23/28) | 83.3 (10/12) | 81.3 (13/16) | 0.643 |
| Less discharge | 73.9 (17/23) | 90.0 (9/10) | 61.5 (8/13) | 0.145 |
| Change in colour | 56.5 (13/23) | 70.0 (7/10) | 46.2 (6/13) | 0.237 |
| Change in smell | 69.6 (16/23) | 80.0 (8/10) | 61.5 (8/13) | 0.313 |

<sup>a</sup>Fisher's exact test was used for the assessment of proportions between groups.

### Testing for the presence of probiotic species in the genital tract

To estimate whether the bacterial species contained in the oral/vaginal probiotic colonised the FGTs of women in the intervention arm, concentrations of *L. rhamnosus, L. acidophilus, B. bifidum* and *B. longum* were measured before and after treatment (Figure 4A). Although not typically considered a vaginal *Lactobacillus* species, it was interesting to note that a third (20/30) of women had detectable concentrations of *L. acidophilus* (median concentration; 6.24×10^−8^ ng/mL) at baseline, although few women had *L. rhamnosus* (2/30), *B. bifidum* (2/30) and *B. longum* (8/30). Despite daily administration of >2×10^9^ CFU vaginally and orally for a total of 15 days in the intervention group, we observed no clear evidence of colonization of any of the bacterial species contained in the probiotic formulation after administration of the probiotic (month 1) or at later time points (Figure 4A).

**Figure 4.**
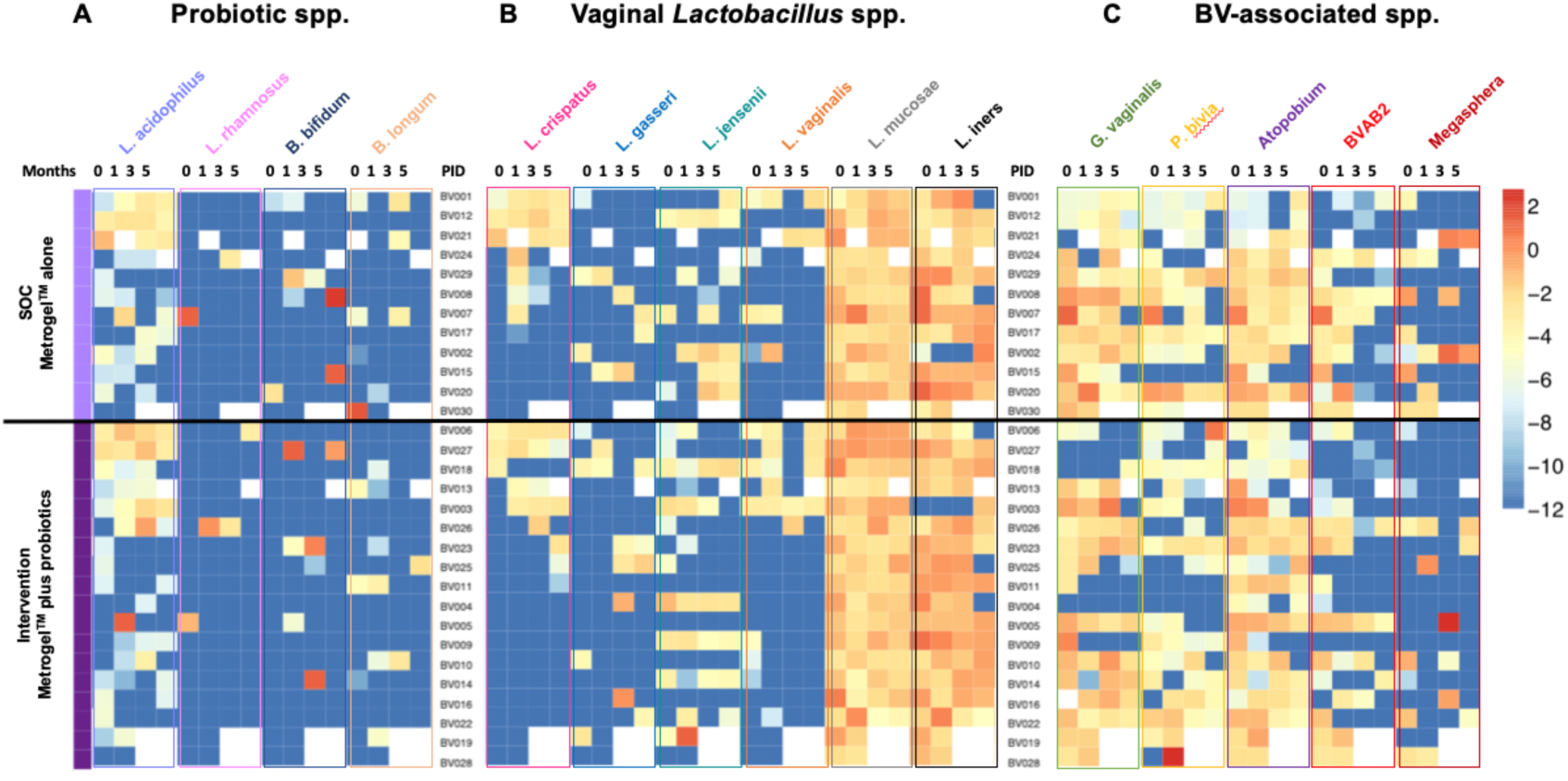
Quantities of vaginal bacterial species in the SOC and intervention group. (A) Bacterial species contained in Vagiforte® Plus Combo Pack, including *L. acidophilus, L. rhamnosus, B. bifidum and B. longum*, vaginal *Lactobacillus spp*., including *L. crispatus, L. gasseri, L. jensenii, L. vaginalis, L. mucosae* and *L. iners* (B) and BV-associated bacteria, *including G. vaginalis, P bivia, Atopobium*, BVAB2 and *Megasphera* (C) were measured by qPCR and normalised to total 16S rRNA gene concentration at month 0 (enrolment) and one, three and five months post-treatment. Values were log-transformed and supervised clustering was used to generate the heatmap. Each row shows one participant. Darkest blue indicates levels below detection limit. White data points indicate missing data. Light purple indicates participants from the SOC and dark purple participants from the intervention group.

### Changes in commensal Lactobacillus communities following BV treatment

*Lactobacillus* spp.-dominated vaginal microbiota are associated with a lack of genital inflammation compared to non-optimal vaginal microbiota (32). However, while some *Lactobacillus* spp., such as *L. crispatus, L. gasseri, L. jensenii* and *L. vaginalis*, have been associated with decreased genital inflammation and HIV risk, the role of others, such as *L. iners*, remains unclear (32,33). Therefore, we evaluated concentrations of several of these *Lactobacillus* spp. to assess the beneficial effect of the oral/vaginal probiotic on the vaginal microbiota. Most women had either concentrations below the detection limit or very low concentrations of *L. crispatus*, *L. gasseri, L. jensenii* and *L. vaginalis* (all with median concentration 0 ng/mL) before treatment (Figure 4B). While some women showed increased concentrations of beneficial vaginal *Lactobacillus* spp. one month after treatment, others did not, and this did not differ by study arm (Additional Table 1). The relative concentrations of *L. mucosae* (median concentration 4.03×10^−2^ ng/mL) and *L. iners* (median concentration 5.23×10^−2^ ng/mL) were generally higher than the concentrations of the other *Lactobacillus* spp. and remained higher over the course of the study (Figure 4B).

**Additional Table 1.** Median vaginal bacterial and IL-1α concentrations in the SOC and intervention arm.

| Median (IQR)<br>[ng/mL] | Pre-treatment |  |  | Month 1 |  |  | Month 3 |  |  | Month 5 |  |  |
| --- | --- | --- | --- | --- | --- | --- | --- | --- | --- | --- | --- | --- |
|  | SOC<br>n=12 | Int.<br>n=18 | p-value | SOC<br>n=11 | Int.<br>n=18 | p-value | SOC<br>n=11 | Int.<br>n=15 | p-value | SOC<br>n=10 | Int.<br>n=14 | p-value |
| *L. crispatus* | 0<br>(0-2.09x10<sup>-6</sup>) | 0<br>(0-0) | 0.2589 | 2.71x10<sup>-6</sup><br>(0-0.001) | 0<br>(0-1.61x10<sup>-5</sup>) | 0.0223 | 0<br>(0-0.003) | 0<br>(0-5.1x10<sup>-5</sup>) | 0.3430 | 0<br>(0-0.0004) | 0<br>(0-0.0005) | 0.2935 |
| *L. gasseri* | 0<br>(0-1.04x10<sup>-7</sup>) | 0<br>(0-5.88x10<sup>-8</sup>) | 0.5310 | 0<br>(0-0) | 0<br>(0-0) | 0.2570 | 0<br>(0-0) | 0<br>(0-9.07x10<sup>-5</sup>) | 0.2957 | 0<br>(0-1.98x10<sup>-5</sup>) | 0<br>(0-2.02x10<sup>-6</sup>) | 0.5138 |
| *L. jensenii* | 0<br>(0-2.71x10<sup>-7</sup>) | 3.79x10<sup>-7</sup><br>(0-0.0004) | 0.0526 | 0<br>(0-1.23x10<sup>-5</sup>) | 6.02x10<sup>-10</sup><br>(0-0.001) | 0.1044 | 1.19x10<sup>-9</sup><br>(0-0.001) | 0<br>(0-2.75x10<sup>-5</sup>) | 0.1149 | 2.65x10<sup>-5</sup><br>(0-0.001) | 0<br>(0-6.21x10<sup>-5</sup>) | 0.0787 |
| *L. vaginalis* | 0<br>(0-3.7x10<sup>-7</sup>) | 0<br>(0-4.45x10<sup>-6</sup>) | 0.4432 | 0<br>(0-0.0001) | 0<br>(0-9.98x10<sup>-5</sup>) | 0.4243 | 0<br>(0-0) | 0<br>(0-0) | 0.4712 | 0<br>(0-0.001) | 0<br>(0-0.001) | 0.5057 |
| *L. mucosae* | 0.024<br>(0.004-0.144) | 0.041<br>(0.006-0.302) | 0.2537 | 0.008<br>(0.002-0.173) | 0.009<br>(0.002-0.020) | 0.4184 | 0.026<br>(0.007-0.122) | 0.023<br>(0.005-0.196) | 0.4349 | 0.020<br>(0.016-0.032) | 0.024<br>(0.008-0.255) | 0.3050 |
| *L. iners* | 0.025<br>(0.001 – 8.345) | 0.067<br>(0.01-0.552) | 0.4668 | 0.024<br>(0.001-0.172) | 0.043<br>(0.007-0.295) | 0.2227 | 0.044<br>(0.013-0.175) | 0.082<br>(0.026-0.256) | 0.3101 | 0.057<br>(0.004-0.456) | 0.069<br>(0.001-0.344) | 0.4887 |
| *L. acidophilus* | 8.79x10<sup>-8</sup><br>(0-8.89x10<sup>-6</sup>) | 5.83x10<sup>-8</sup><br>(0-2.22x10<sup>-6</sup>) | 0.3867 | 2.00x10<sup>-8</sup><br>(0-3.56x10<sup>-5</sup>) | 6.01x10<sup>-9</sup><br>(0-2.53x10<sup>-5</sup>) | 0.4897 | 3.37x10<sup>-8</sup><br>(0-0.0004) | 2.23x10<sup>-7</sup><br>(0-0.010) | 0.3918 | 6.86x10<sup>-7</sup><br>(0-0.0001) | 1.85x10<sup>-7</sup><br>(0-0.0001) | 0.4123 |
| *L. rhamnosus* | 0<br>(0-0) | 0<br>(0-0) | 0.4000 | 0<br>(0-0) | 0<br>(0-0) | 0.6207 | 0<br>(0-0) | 0<br>(0-0) | 0.6769 | 0<br>(0-0) | 0<br>(0-0) | 0.5833 |
| *B. bifidum* | 0<br>(0-0) | 0<br>(0-0) | 0.1517 | 0<br>(0-4.40x10<sup>-9</sup>) | 0<br>(0-0) | 0.3799 | 0<br>(0-0) | 0<br>(0-0) | 0.2587 | 0<br>(0-6.735) | 0<br>(0-0) | 0.1630 |
| *B. longum* | 0<br>(0-1.19x10<sup>-6</sup>) | 0<br>(0-0) | 0.1021 | 0<br>(0-0) | 0<br>(0-5.61x10<sup>-8</sup>) | 0.0578 | 0<br>(0-7.21x10<sup>-5</sup>) | 0<br>(0-0) | 0.2403 | 0<br>(0-0) | 0<br>(0-0) | 0.5833 |
| *G. vaginalis* | 0.025<br>(0.0001 – 0.358) | 0.001<br>(7.6x10<sup>-6</sup>-0.067) | 0.2069 | 0.001<br>(7.75x10<sup>-6</sup>-0.018) | 0.002<br>(0-0.017) | 0.2774 | 0.001<br>(9.38x10<sup>-5</sup>-0.05) | 0.001<br>(0-0.156) | 0.3210 | 0.0003<br>(3.01x10<sup>-8</sup>-0.006) | 2.91x10<sup>-5</sup><br>(0-0.016) | 0.4577 |
| *P. bivia* | 2.78x10<sup>-5</sup><br>(5.11x10<sup>-7</sup> -0.015) | 4.51x10<sup>-6</sup><br>(0-0.0001) | 0.0969 | 2.99x10<sup>-6</sup><br>(0-3.15x10<sup>-5</sup>) | 1.72x10<sup>-5</sup><br>(0-0.002) | 0.3518 | 1.05x10<sup>-5</sup><br>(3.75x10<sup>-7</sup>-0.0002) | 1.1x10<sup>-6</sup><br>(0-1.48x10<sup>-5</sup>) | 0.1232 | 0<br>(0-0.002) | 5.36x10<sup>-6</sup><br>(0-6.30x10<sup>-5</sup>) | 0.3847 |
| *Atopobium* | 0.009<br>(0.001-0.225) | 0.008<br>(1.2x10<sup>-5</sup>-0.1) | 0.4881 | 0.001<br>(3.54x10<sup>-6</sup>-0.005) | 0.003<br>(1.04x10<sup>-6</sup>-0.057) | 0.3299 | 0.002<br>(0-0.014) | 1.23x10<sup>-5</sup><br>(3.97x10<sup>-10</sup>-0.019) | 0.3447 | 0.0001<br>(6.02x10<sup>-6</sup>-0.003) | 0.0003<br>(0-0.003) | 0.4367 |
| *BVAB2* | 0.0005<br>(1.54x10<sup>-7</sup>-0.006) | 4.51x10<sup>-7</sup><br>(0-0.0001) | 0.2739 | 4.68x10<sup>-5</sup><br>(0-0.002) | 9.06x10<sup>-7</sup><br>(0-0.002) | 0.2261 | 6.26x10<sup>-11</sup><br>(0-3.34x10<sup>-5</sup>) | 0<br>(0-6.65x10<sup>-10</sup>) | 0.2590 | 0<br>(0-8.07x10<sup>-7</sup>) | 0<br>(0-1.05x10<sup>-5</sup>) | 0.4103 |
| *Megasphaera* | 5.53x10<sup>-5</sup><br>(0-0.003) | 0<br>(0-3.28x10<sup>-5</sup>) | 0.0922 | 0<br>(0-0.0002) | 0<br>(0-0) | 0.2036 | 0<br>(0-0.108) | 0<br>(0-0) | 0.3457 | 0<br>(0-0.172) | 0<br>(0-0) | 0.2421 |
| IL-1α [pg/mL] | 4.80<br>(0.64-37.63) | 11.26<br>(0.64-19.16) | 0.4044 | 1.28<br>(0.64-35.84) | 10.42<br>(0.64-32.18) | 0.1918 | 4.36<br>(0.64-12.74) | 11.96<br>(2.68-19.33) | 0.0491 | 6.37<br>(3.04-33.03) | 9.57<br>(0.64-49.65) | 0.4622 |

The Mann-Whitney test was used to compare SOC and intervention arms.

### Changes in BV-associated bacterial communities following BV treatment

Most women had detectable concentrations of *G. vaginalis* (26/30; median concentration 1.25×10^−2^ ng/mL), *P. bivia* (21/30; median concentration 7.82×10^−3^ ng/mL) and *A. vaginae* (29/30; median concentration 1.22×10^−5^ ng/mL) pre-treatment (Figure 4C), consistent with their BV status. The concentrations of these BV-associated bacteria remained high throughout the trial despite BV treatment, both in the SOC and intervention arm (Additional Table 1). While most women also had BVAB2 (22/30; 3.31×10^−5^ and some *Megasphera* (14/30) present pre-treatment, some women experienced a loss or decreased quantities of these bacterial species post-treatment, but this was again irrespective of study arm (Figure 4C).

### Changes in genital cytokine concentrations following treatment

IL-1α concentrations were measured in vaginal samples collected pre- and post-treatment to investigate whether probiotics influenced genital inflammatory cytokine concentrations compared to SOC. Overall, we found that genital IL-1α concentrations decreased after treatment (from a median concentration of 4.80 pg/mL to 1.28 pg/mL in SOC participants vs. from 11.26 pg/mL to 10.42 pg/mL in the intervention group; Figure 5; Additional Table 1), although not significantly. In addition, adjusting for STIs that were acquired during the trial had no impact on this observation.

**Figure 5.**
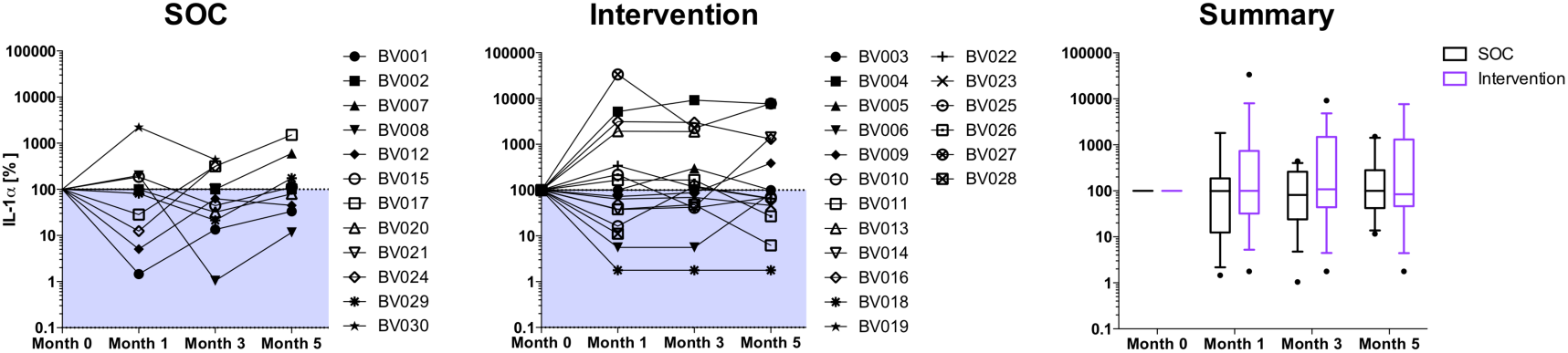
Levels of the inflammatory marker IL-1α in the FGT of participants in the SOC and intervention arm. IL-1α was measured by ELISA in FGT secretions of SOC and intervention group participants pre-treatment and 1, 3 and 5 months post-treatment. Each participant is represented by a symbol-coded dot. The percentages compared to the IL-1α level measured pre-treatment (month 0) are displayed. The shaded area indicates a decrease in IL-1α post-treatment. The summary shows the median and interquartile range.

## Discussion

We aimed to explore the regulatory environment in South Africa by conducting a clinical trial with a commercially available OTC probiotic, and thereby to lay the groundwork for assessing the effect of adjunctive probiotics on BV treatment in South Africa. The regulatory environment in South Africa continues to consider OTC probiotics as health supplements, and SAHPRA formally approved this trial. We found that the use of a vaginal/oral probiotic combination was well accepted among South African women and associated with few AEs. Despite general acceptance of the product, we found that this particular oral/vaginal probiotic did not significantly improve BV cure rates following antibiotic treatment of symptomatic BV, neither did it lower vaginal pH or improve BV recurrence within five months post-treatment compared to MetroGel™ alone (SOC), although it needs to be emphasises that our sample was not powered to detect anything other than large changes. This result, while disappointing, also calls for stricter regulations of probiotics in South Africa. The World Health Organization defines probiotics as “live microorganisms which when administered in adequate amounts confer a health benefit on the host”, thus in order to be labelled a probiotic, scientific evidence demonstrating a health benefit needs to be well documented. Products that contain bacterial strains with no proven health benefit should not be labelled probiotics and marketed as such, as these strains, strictly spoken, cannot be considered probiotics as the health benefit is included in the definition of a probiotic. Marketing “probiotics” with no proven health benefit may lead to misleading perceptions and generalised criticism of all probiotics, although some do have a proven health benefit.

While we were not able to detect a beneficial effect of adjunctive Vagiforte® PLUS Combo Pack under these trial conditions, possibly due to the small sample size, the product was found to be generally safe. Women reportedly preferred the oral over the vaginal application of the probiotic, as the administration of the vaginal spray (in its reusable spray applicator) was uncomfortable and messy. Thus, for future probiotic product design, women’s preferences should be taken into consideration and probiotics containing well-selected vaginal *Lactobacillus* spp. should be administered orally or topically as vaginal tablet or gel. Importantly, women were not willing to spend more than ZAR 200 for one treatment course. Considering the current prices for probiotics on the South African market (22), this emphasizes the need for affordable treatment options for those most in need of it.

Ideally, the vaginal microbiota should be dominated by beneficial *Lactobacillus* spp. (32), thereby contributing to the effectiveness of the host mucosal barrier against infection. The selected OTC oral/vaginal probiotic, like all those available in South Africa, did not contain *Lactobacillus* spp. that are commonly associated with vaginal health, internationally and in South African women (19). It was therefore not unexpected that both the proportion of women and the relative concentrations of Vagiforte^®^ bacterial species were found to be similar in women in the intervention and SOC arm, and evidence of probiotic colonization was lacking. This suggests that the probiotic strains did not effectively colonize the FGTs of women randomised to the intervention arm, despite having proven viability at the initiation of the trial. This may be due to the fact that Vagiforte^®^ PLUS did not contain vaginal *Lactobacillus* spp. commonly associated with reproductive health (such as *L. crispatus, L. gasseri, L. jensenii* or *L. vaginalis)* and thus likely did not effectively colonise the FGT. Similarly, others have reported that supplementation of standard antibiotic therapy with commercially-available probiotic lactobacilli in South African women with BV does not confer a measurable benefit (18), indicating that better designed probiotic products for treating BV need to be developed and made available for South African women. These products should contain well characterised vaginal *Lactobacillus* strains with confirmed beneficial probiotic characteristics that have ideally been isolated from the FGTs of healthy South African women.

Some additional recommendations for the design of future probiotic trials in South Africa can be made from this study. Clinical studies should test for BV cure shortly after treatment completion to minimize the likelihood that women have a recurrent BV event, in order to differentiate BV cure rates from confounding repeat episodes. Further, this trial was a superiority trial rather than placebo-controlled, as we tested the intervention against MetroGel™ instead of placebo or oral metronidazole as control group (34), which is likely to have made it harder to detect differences. Further, oral metronidazole is in fact more commonly used in South Africa for treating BV due to its lower cost compared to MetroGel™, which is slightly more effective and has less systemic side effects than the cheaper oral formulation (35,36).

The limitations of this study include the small number of participants, however this was a pilot intended to test the regulatory environment. Further, administration of the study drugs was self-reported and vaginal pH was not being measured pre-treatment. Further, no 16S rRNA gene sequencing to evaluate relative vaginal bacterial abundance was performed. However, detailed information on vaginal practices, behavioural and sexual risk factors, study product use, side effects, and concomitant medications were collected which allowed assessment of potential confounders.

## Conclusions

This randomized single-blinded trial provided important regulatory and clinical considerations critical in the design of future clinical trial, with improved probiotic products and larder sample siezes. Although the trial was not powered to determine a beneficial effect of this probiotic on BV cure in South African women over metronidazole treatment alone, it showed that oral and vaginal probiotics were generally safe to use and acceptable in South African BV positive women. Due to the critical need to manage BV better internationally and in South Africa, future regulatory approved, double-blind, randomized, placebo-controlled trial powered for efficacy, using a probiotic product containing beneficial, well-characterised vaginal *Lactobacillus* strains are needed to conclusively determine the efficacy of adjunctive probiotics on BV cure and recurrence in South African women. In a region with some of the highest rates of BV, STIs and HIV, it is critical that the regulatory environment is established and tested to facilitate the evaluation of these improved products once they enter clinical trials.

**List of Abbreviations**
BV: - Bacterial vaginosis
BVAB2: - Bacterial Vaginosis-Associated Bacterium 2
CFU: - colony-forming units
CRC: - Clinical Research Centre
FGT: - female genital tract
ICH: - International Conference on Harmonisation
OTC: - over-the-counter
RCT: - randomized controlled trial
SA-GCP: - South African Good Clinical Practice
SAHPRA: - South African Health Products Regulatory Authority
sBHI: - Brain Heart Infusion supplemented with 0.1% starch and 1% yeast
SOC: - Standard of care
STI: - sexually transmitted infections
UCT: - University of Cape Town
ZAR: - South African Rand

## Data Availability

The datasets used and/or analysed during the current study are available from the corresponding author on reasonable request.

## Declarations

### Ethics approval and consent to participate

Approval for this clinical trial was obtained from University of Cape Town (UCT) Human Research Ethics Committee (HREC Ref 706/2016) and the South African National Health Research Ethics Council (NHREC Ref 4579). The trial was registered with the South African National Clinical Trial Register of the Department of Health (DOH-27-1117-5579; PI: S. Barnabas) and conducted in full compliance with South African Good Clinical Practice (SA-GCP) and International Conference on Harmonisation (ICH)-GCP, supported by the UCT Clinical Research Centre (CRC). Before undergoing any trial-related procedures, all women provided written informed consent.

### Consent for publication

Not applicable.

### Authors’ Contributions

AUH designed the trial, enrolled the cohort, performed the experiments, analysis and wrote the manuscript. RS and NM performed some of the experiments. HJ and KM contributed to writing the paper. SLB and JSP designed the trial, enrolled the cohort and contributed to analysis and writing the paper. All authors have read and approved the manuscript.

## Acknowledgements

We would like to thank the UCT CRC for their regulatory support and help in setting up and running the trial.

